# Automated Oxygen Delivery in Home Setting for Patients with COPD on Long-Term Oxygen Therapy – a randomized crossover feasibility trial

**DOI:** 10.1101/2025.01.23.25320958

**Authors:** Linette Marie Kofod, Ejvind Frausing Hansen, Morten Tange Kristensen, Barbara Cristina Brocki, Elisabeth Westerdahl

## Abstract

**Rationale:** Patients with COPD on long-term oxygen therapy (LTOT) have an unmet need for oxygen adjustments during sleep, rest, and activity, documented by continuous monitoring of oxygen saturation. While emerging technology enables automated adjustments, its feasibility in home settings remains uncertain. This randomized crossover trial evaluated the feasibility and preliminary effects of continuous automated oxygen titration in the homes of patients on LTOT.

**Methods:** The intervention involved four days of automated oxygen titration targeting a SpO₂ of 90-94% using a Bluetooth-connected electronic device and wrist pulse oximeter, forming a closed-loop system. Oxygen flow (0.9-6.8 L/min) was continuously adjusted based on SpO₂. During the control period, patients received their usual fixed dose oxygen. Feasibility was defined as successful automated titration time and time spent with normoxia. Changes in health status were measured using the Clinical COPD Questionnaire (CCQ).

**Results:** Twelve patients on LTOT (2.0±0.8 L/min) were included, with more than 217,000 paired SpO_2_ and oxygen flow data points collected per patient. Oxygen flow was automatically adjusted for a median (IQR) of 77 (68.0–84.3) hours, covering 83% of the time. Time within target saturation increased significantly from 52% (42–63) to 86% (75–90) during intervention, with all patients utilizing the full available flow range. The CCQ score improved by 0.74±0.47 points, p <0.001.

**Conclusion:** Automated home oxygen titration is feasible, achieving more time with normoxia, but it required a wide flow range and continuous monitoring. The patients reported notable reductions in COPD symptoms.

## Introduction

Oxygen is essential for human life. Regrettably, some patients with advanced COPD develop a reduced ability to deliver sufficient oxygen to the blood, resulting in chronic respiratory failure with the constant need of oxygen supplementation.

In acute settings, oxygen treatment has become more restrictive, with increased focus on the dosage of oxygen for patients with acute hypoxemic respiratory failure, and it is recommended to maintain a target saturation with conservative oxygen doses, as it decreases mortality [1–3]. Several studies found that electronic, closed-loop devices, which automatically adjusts oxygen in response to the saturation, were more accurate in keeping the recommended saturation compared to manual adjustments [4–8].

In contrast, our understanding is limited when it comes to maintaining target saturation during daily living in the home setting. Fluctuating oxygen needs lead to episodes of desaturation and hypoxemia in patients with COPD [9–13]. Home oxygen is prescribed based on arterial blood gas analysis to achieve a PaO2 >8.0 kPa, corresponding to a peripheral oxygen saturation (SpO_2_) of 90% at rest [14,15]. Although patients on long-term oxygen therapy (LTOT) use this prescribed fixed oxygen dose, continuous monitoring of oxygen saturation has revealed an unmet need for oxygen adjustments, depending on whether the patients are sleeping, sitting, or engaging in daily activities [11,16]. Clinical benefits of maintaining a target saturation are particular evident during walking tests, with increased walking capacity and alleviated dyspnea [17–19], but also when performing an activity of daily living (ADL) test [20]. Optimizing patients’ oxygen saturation through automated oxygen titration enhanced their ability to perform ADL and reduced their perceived breathlessness [20]. These results suggest that individually adjusted, automated oxygen flow may benefit patients in daily life by increasing time spent with normoxia. However, achieving this would require constant saturation monitoring around the clock and, consequently, continuous adjustments.

Automated oxygen titration technology has advanced into the home setting, allowing closed-loop devices to be attached to the patients’ home concentrators. For oxygen flow to dynamically adjust in every situation, patients need to wear a pulse oximeter that communicates with the oxygen delivery system for most of the day. This represents a new approach, and it remains unclear how, or even if, such oxygen titration could be effectively managed.

The aim of this randomized crossover study was to evaluate the feasibility of automated oxygen titration as response to the saturation during daily living for four days in the home of patients with COPD on LTOT. Feasibility was defined as successful time during which the patients were automatically titrated, patientś willingness toward the intervention, and clinically relevance.

## Methods

### Study design

This randomized crossover feasibility trial was conducted in the homes of 12 patients with COPD on LTOT. The patients were recruited from two departments of pulmonology at Copenhagen University Hospital, Hvidovre and Copenhagen University Hospital, Bispebjerg-Frederiksberg, Denmark, from January to December 2023 in connection with a scheduled study visit [20].

The study was approved by The Committees on Health Research Ethics in the Capital Region of Denmark (H-22032988) and the Danish Data Protection Agency j.nr. P-2022-625. The study was registered at ClinicalTrials.gov (NCT05556187), and the reporting followed the CONSORT statement for randomized pilot and feasibility trials.

### Participants

≤7.3 kPa), who were receiving LTOT according to the international criteria for home oxygen therapy [14], had the ability to walk independently (with or without a walking aid), and were cognitively able to participate. Exclusion criteria were an exacerbation in COPD treated with either antibiotics or prednisolone within the preceding three weeks or comorbidities known to impact physical functioning. Additionally, before randomization, each patient underwent two venous blood gas tests: the first with their usual oxygen dose and the second after 20 minutes of 8 L/min of oxygen. Venous blood gases (instead of arterial blood gases) were used to minimize discomfort to the patients [21]. The blood samples were analyzed for pH-value and PvCO_2_. The patients were excluded if they exhibited a drop in pH to <7.31 on 8 L/min of oxygen flow or an increase in the partial pressure of carbon dioxide within venous blood (PvCO_2_) of >1 kPa compared to their usual fixed oxygen dose.

Included patients provided written informed consent before participation.

### Intervention

#### The automated oxygen period

The intervention period consisted of four days with a continuous titration of the oxygen flow aiming at a target saturation between 90-94% using an electronic closed-loop device installed in the patientś homes. If SpO_2_ dropped below 90% or exceeded 94% the oxygen flow was automatically adjusted according to the algorithm in the device.

#### The fixed dose period

For the control period, the patients received their usual fixed oxygen flow for four days, while monitoring and collecting data on SpO_2_ and heart rate.

The two periods were scheduled on comparable days to minimize significant variations in social or physical activities; however, no specific washout time was planned. In both periods, daily 24-hour physical activity were monitored with a body-worn device, and the patients used their usual portable oxygen concentrator when being outdoors.

### Study technology and data capture

#### The automated oxygen period

A closed-loop device O2matic Home Oxygen Treatment (HOT) (O2matic Ltd., Herlev, Denmark) was used for automated oxygen titration. The 9 L/min Invacare Platinum 9 concentrator (Invacare Ltd. Brøndby, Denmark) or the 10 L/min Caire NewLife Intensity concentrator (Medical Danmark Ltd., Ejby, Denmark) were used to produce oxygen and attached to the HOT device using a tube.

The patients wore the Nonin Wrist Pulse Oximeter (Nonin Medical, Inc., USA) with the sensor well-attached to a finger using a sensor tape, Figure 1. The pulse oximeter transmitted data on SpO_2_ and heart rate to the HOT device. The adjustments on oxygen were done every second based on average SpO_2_ for the last 15 seconds. All parameters regarding the individual patient settings could only be changed in the Installer tablet handled by the investigator. The SpO_2_ target interval and a flow range was set at the beginning of the intervention. A previous study identified an mean oxygen demand of 8 L/min during a walking test [19].

**Figure 1.**
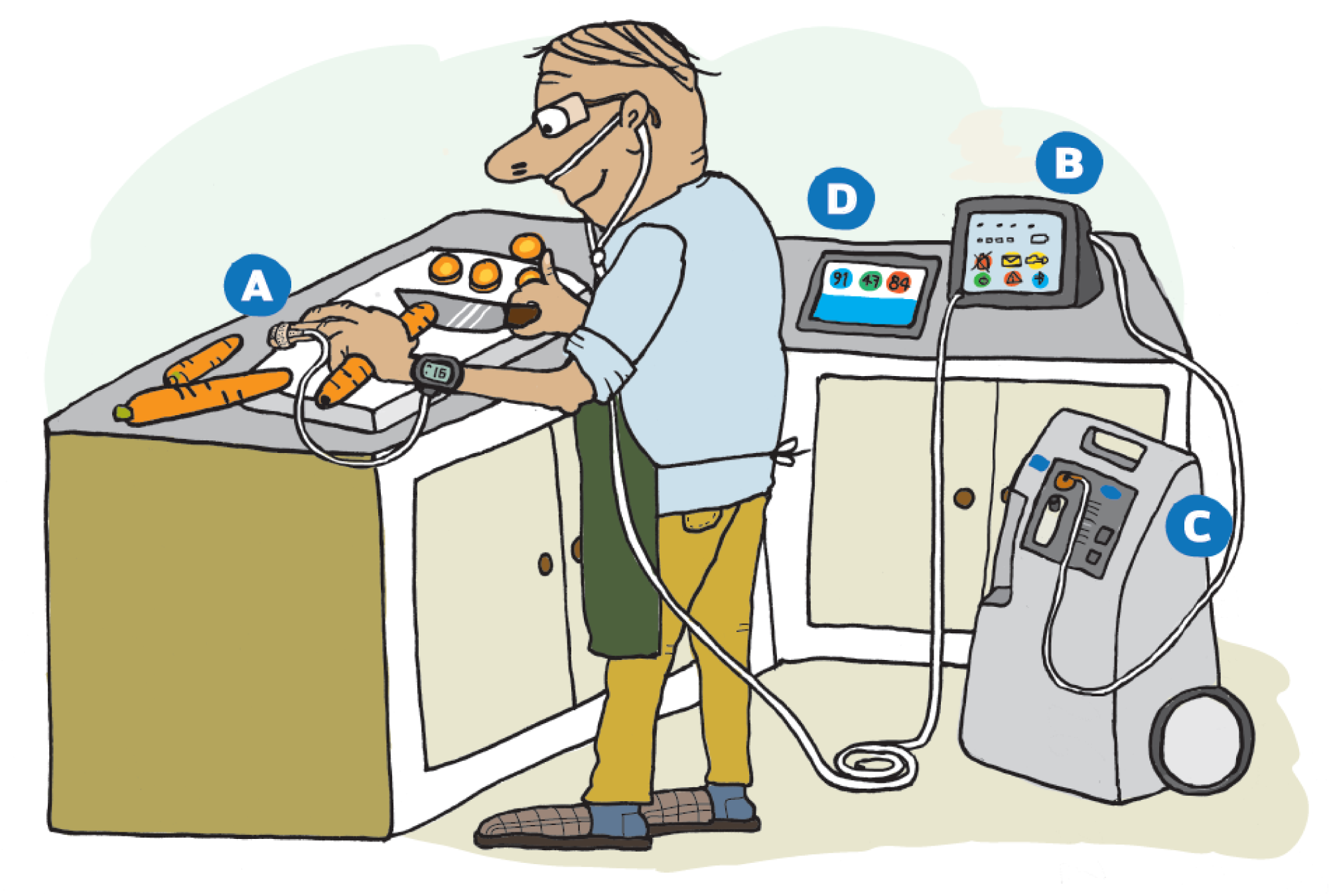
Artistic presentation of the automated oxygen delivery setup. A) Pulse oximeter, which transmits oxygen saturation data via Bluetooth to the closed-loop devise, B) Closed-loop device, that titrates oxygen via the high-flow nasal cannula to the patient, C) Oxygen concentrator, D) Patient tablet, displaying the current saturation, oxygen flow and heart rate. The patient tablet also transmits information via Wi-fi to the online platform.

Therefore, the aim was an oxygen flow from 0.5 to 8.0 L/min. However, the concentrators were only able to deliver a stable flow between 0.8 and 6.8 L/min. In case the patients removed the pulse oximeter (or experienced a general loss of signal) for more than two minutes the flow would return to usual flow ±1 L/min and stay there until the signal was restored.

#### Data collection

The close-loop device transmitted data to an app on the patient-tablet. The data were then relayed via Wi-Fi to a secure web platform based on Microsoft Azure cloud services, where it could be accessed by the principal investigator as needed. The patients could also observe live data on the tablet if they wished.

In case of a disconnection to the patient-tablet, data were stored locally on the closed-loop device and transmitted once the connection was restored. During such interruptions, the oxygen flow to the patient remained automated titrated, and the stored data could later be accessed on the online platform as summarized, non-live data files. Live data refers to every-second-real-time measurements of heart rate, SpO_2_, and oxygen flow, which could be extracted from CSV files.

#### The fixed dose period

During the fixed dose period, the patients wore the same wrist pulse oximeter as during the intervention. Since automated oxygen titration could only be administered in the patient’s home, we instructed the patients to remove the pulse oximeters when leaving the house during the control period, so that data was only collected when being indoors.

#### Data collection

The data on SpO_2_ and heart rate were stored locally in the wrist oximeter. CSV files, used for analysis, was generated using Nonin’s nVISION Data Management Software. The batteries in the pulse oximeter needed to be changed every second day, and when removed, it resulted in a reset of date and time in the system. As a result, the collected data during the fixed dose period was untransparent, regarding the exact date and time.

#### Step counts

The physical activity was monitored equally in both study periods using the SENS motion accelerometer (SENS innovation ApS, Denmark). The miniature, waterproof accelerometer was placed with a patch above the patientś knee. It continuously recorded the movements, and synchronized with an app that uploaded the recorded data to a web server [22]. Summary files containing information on steps were accessed from SENS online platform and used for analysis.

### Outcome

#### Feasibility

The goal of this study was for patients to wear the pulse oximeter continuously during the daytime while having their oxygen adjusted automatically. We defined three key areas that needed to be fulfilled for it to be considered feasible: time when the patients were automatically titrated, patientś willingness toward the intervention and clinically relevance. Prior to the study, we established the following criteria:

1. Data were successfully transmitted from the wrist pulse oximeter to the closed-loop device and further to the cloud solution (<10 % data loss),
2. The patients wore the wrist pulse oximeter for more than 50% of the daytime (08:00-20:00),
3. The time spent within target saturation was statistically different between arms and in favour of automated oxygen titration with a difference of at least 10%,
4. The patients were at least as active with automated oxygen titration as with conventional fixed oxygen therapy, measured by the activity sensor,
5. The patients were safe with no serious adverse events, leading to unscheduled healthcare contacts.

Rationale for criteria 1, *Titration time*: Both live data and session summaries received on the platform were proof of “the time when the patients were automatically titrated”. The 10% threshold was based on experiences from three earlier studies on automated oxygen delivery [7,19,20].

Rationale for criteria 2, *Patient willingness*: We considered patients’ willingness to wear the wrist pulse oximeter in the day time from 8 a.m. to 8 p.m. as essential for continuous oxygen titration.

Rationale for criteria 3 and 4, *Clinical relevance*: If automated oxygen titration did not improve oxygen saturation, it was deemed irrelevant. Therefore, we evaluated: differences in time spent with normoxia (SpO_2_ 90–94%), moderate hypoxemia (SpO_2_ 85-89%), severe hypoxemia (SpO_2_<85%) and hyperoxemia (SpO_2_>94%). Furthermore, we evaluated the patients’ physical activity level.

Rationale for criteria 5, *Patient safety:* The setup was considered non-feasible if it posed notable risks to the patient, such as unscheduled contacts to the hospital due to acute-on-chronic respiratory failure.

#### Secondary outcome

At study start, after four days intervention, and after four days of usual care, the patients were assessed using the health status Clinical COPD Questionnaire (CCQ), 24 hour version [23]. The CCQ is validated in Danish, consists of ten questions across three domains: symptoms, mental state, and functional state. Each question is scored from 0 to 6, with a higher score indicating a lower health status. A Minimal Important Difference (MID) of 0.4 is considered clinically relevant [24].

The patients’ oxygen flow was compared between both study periods.

#### Variables

We included the following variables to describe the characteristics and clinical profile of the patients: age, gender, body mass index (BMI), pulmonary function and duration of LTOT, all of which were extracted from the patients’ medical records. Additionally, the patients were asked about their perceived dyspnea at rest, measured using the Borg CR10 dyspnea scale, their marital status, need for a walking aid, and frequency of engaging in activities outside their home. The patients also completed the modified Medical Research Council Dyspnea Scale (mMRC) and the COPD Assessment Test (CAT).

### Randomization and blinding

The patients were randomized after inclusion to either the automated oxygen period followed by the fixed dose period, or vice versa. The randomization list was computer-generated compiled for each patient in REDCap electronic data capture tools (REDCap Consortium, Nashville, US) hosted at Capital Region of Denmark.

Neither the patients nor the investigator were blinded.

### Statistical considerations

Due to the feasibility design of the present study, a formal sample size calculation was not performed. In a non-feasibility design CCQ could be a primary outcome, which would require 42 patients to detect a MID of 0.4 with a standard deviation of 0.9. For the purpose of this feasibility study, we selected a sample of 12 patients, as we considered this number sufficient to assess feasibility. Furthermore, this sample size provided an adequate basis for evaluating variations in SpO_2_ intervals and generating preliminary insights into the CCQ outcome.

For the time-spent-within-target-analysis in this study, only *live data* extracted from CSV files were used. In Criteria 1, however, live data were supplemented with the duration of the non-live data to provide a complete picture of the ‘titration tim’.

Continuous variables were examined for normality. Those meeting normality assumptions were analyzed with a paired t-test and presented as mean (standard deviation (SD)). For not normally distributed variables, the Wilcoxon signed-rank test was used, with data presented as median with interquartile range (IQR). CCQ was tested in a two-way ANOVA with treatment, period and an interaction term between treatment and period as explanatory variables to account for carryover effect bias. IBM SPSS Statistics for Windows, ver. 29.01 was used for all statistical analyses. GraphPad Prism version 10.1.2 for Windows was used for figure 3 and 4.

## Results

Thirty-two patients were screened for eligibility, of whom 13 met the inclusion criteria and consented to participate, Figure 2. One patient was subsequently excluded after venous blood gas analysis revealed a PCO₂ of 11.3 kPa and a pH decrease from 7.30 to 7.28 following 20 minutes with 8 L/min oxygen. Accordingly, 12 patients (four women and eight men) with a mean (SD) home oxygen dose of 2.0 (0.7) L/min were randomized, Table 1. All patients completed the study.

**Figure 2.**
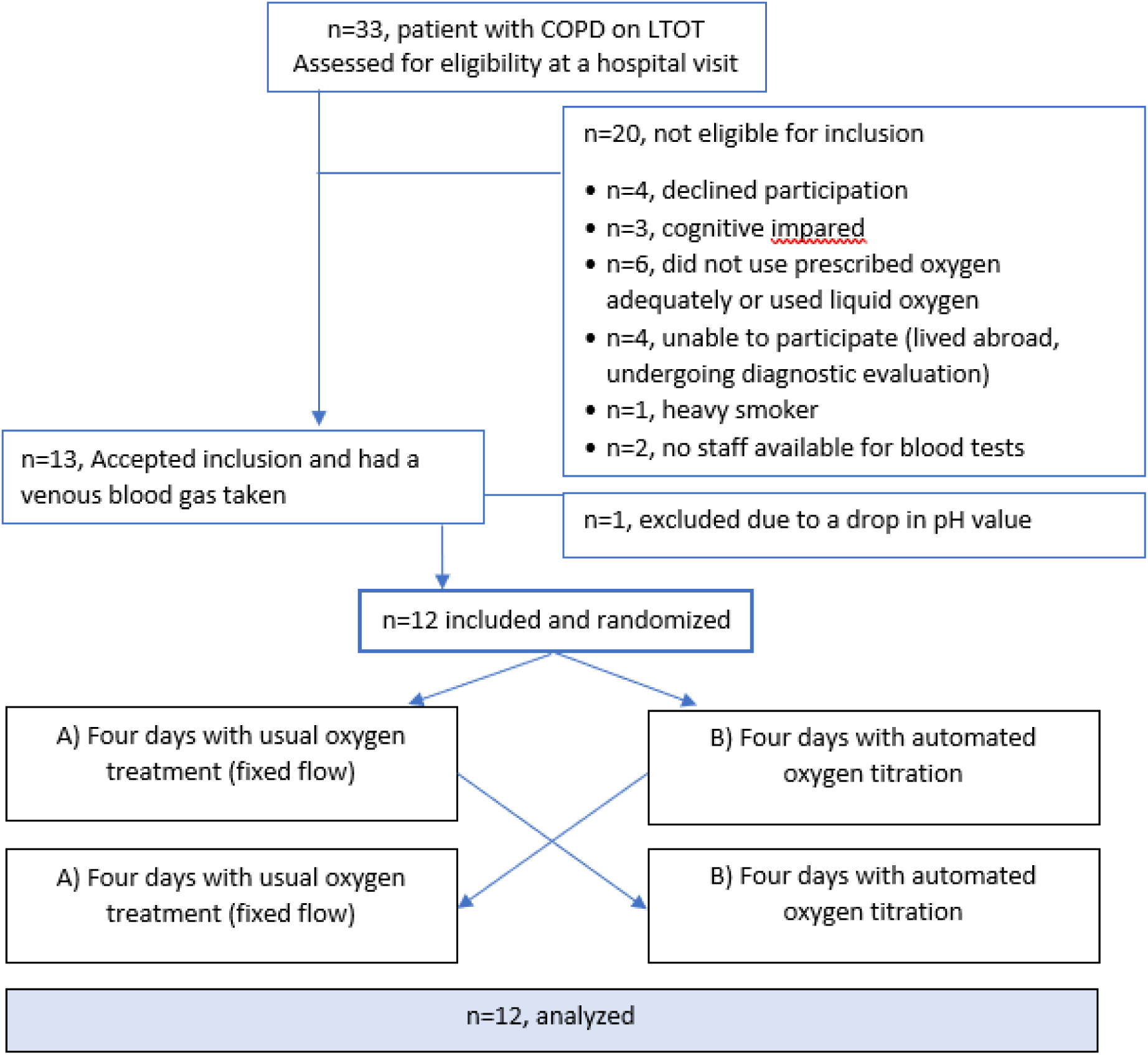
Flow diagram of inclusion of patients.

**Table 1.**
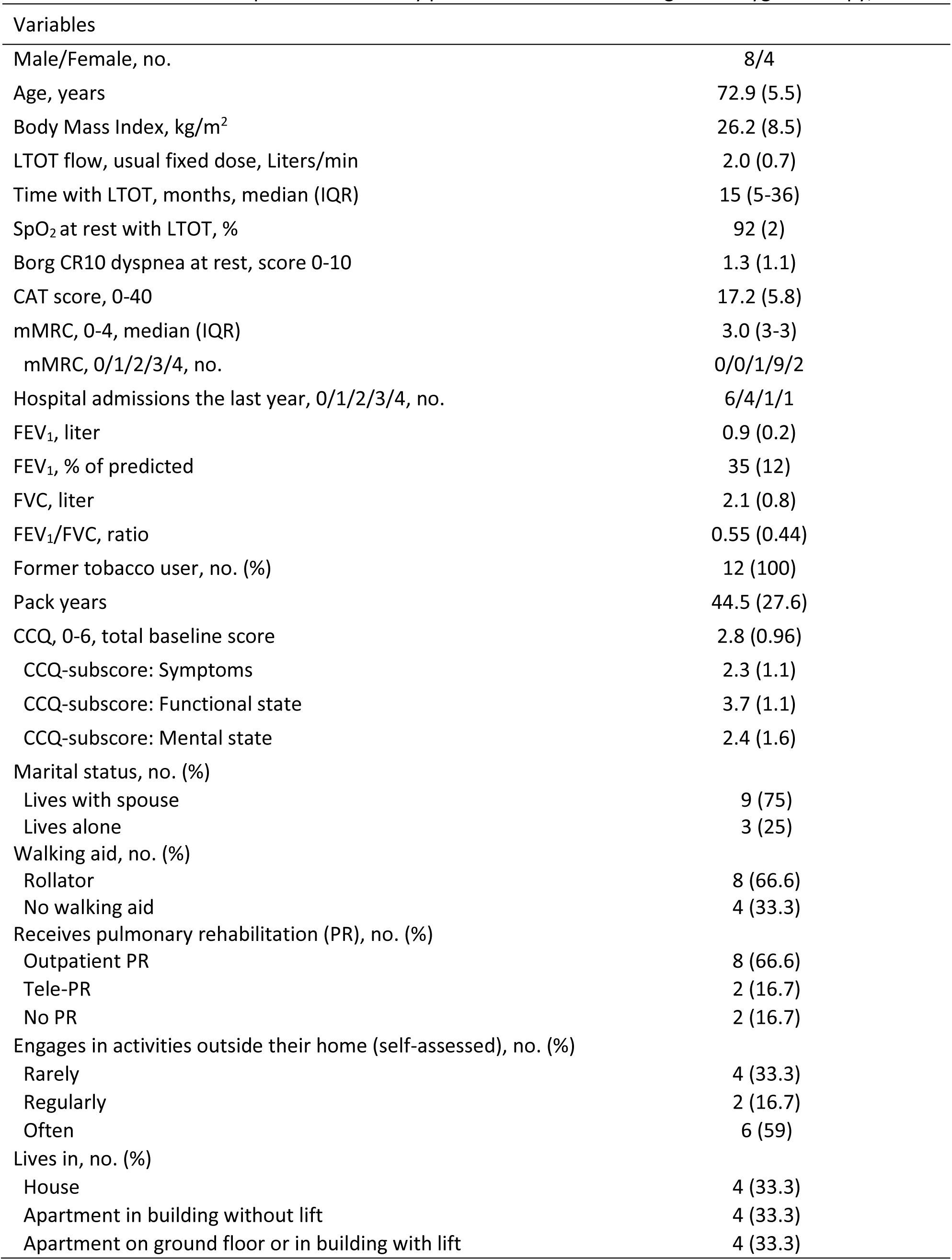
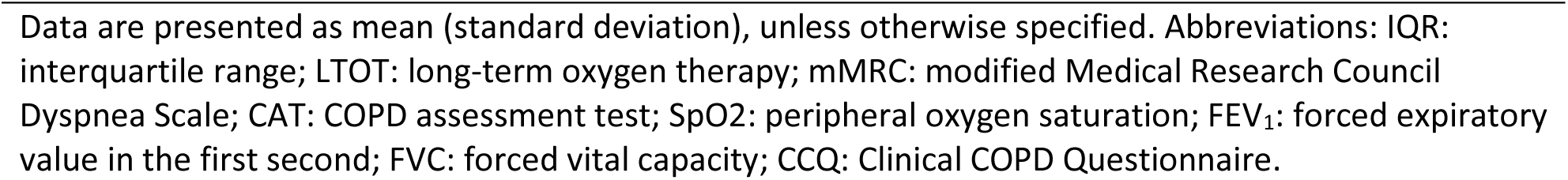
Characteristics and clinical profile of the study patients with COPD on long-term oxygen therapy, n=12.

| Variables |  |
| --- | --- |
| Male/Female, no. | 8/4 |
| Age, years | 72.9 (5.5) |
| Body Mass Index, kg/m<sup>2</sup> | 26.2 (8.5) |
| LTOT flow, usual fixed dose, Liters/min | 2.0 (0.7) |
| Time with LTOT, months, median (IQR) | 15 (5-36) |
| SpO<sub>2</sub> at rest with LTOT, % | 92 (2) |
| Borg CR10 dyspnea at rest, score 0-10 | 1.3 (1.1) |
| CAT score, 0-40 | 17.2 (5.8) |
| mMRC, 0-4, median (IQR) | 3.0 (3-3) |
| mMRC, 0/1/2/3/4, no. | 0/0/1/9/2 |
| Hospital admissions the last year, 0/1/2/3/4, no. | 6/4/1/1 |
| FEV<sub>1</sub>, liter | 0.9 (0.2) |
| FEV<sub>1</sub>, % of predicted | 35 (12) |
| FVC, liter | 2.1 (0.8) |
| FEV<sub>1</sub>/FVC, ratio | 0.55 (0.44) |
| Former tobacco user, no. (%) | 12 (100) |
| Pack years | 44.5 (27.6) |
| CCQ, 0-6, total baseline score | 2.8 (0.96) |
| CCQ-subscore: Symptoms | 2.3 (1.1) |
| CCQ-subscore: Functional state | 3.7 (1.1) |
| CCQ-subscore: Mental state | 2.4 (1.6) |
| Marital status, no. (%) |  |
| Lives with spouse | 9 (75) |
| Lives alone | 3 (25) |
| Walking aid, no. (%) |  |
| Rollator | 8 (66.6) |
| No walking aid | 4 (33.3) |
| Receives pulmonary rehabilitation (PR), no. (%) |  |
| Outpatient PR | 8 (66.6) |
| Tele-PR | 2 (16.7) |
| No PR | 2 (16.7) |
| Engages in activities outside their home (self-assessed), no. (%) |  |
| Rarely | 4 (33.3) |
| Regularly | 2 (16.7) |
| Often | 6 (59) |
| Lives in, no. (%) |  |
| House | 4 (33.3) |
| Apartment in building without lift | 4 (33.3) |
| Apartment on ground floor or in building with lift | 4 (33.3) |

Data are presented as mean (standard deviation), unless otherwise specified. Abbreviations: IQR: interquartile range; LTOT: long-term oxygen therapy; mMRC: modified Medical Research Council Dyspnea Scale; CAT: COPD assessment test; SpO<sub>2</sub>: peripheral oxygen saturation; FEV<sub>1</sub>: forced expiratory value in the first second; FVC: forced vital capacity; CCQ: Clinical COPD Questionnaire.

### Feasibility

During the four days of automated oxygen intervention, the patients had the equipment installed for median (IQR) 96 (94.7-96.0) hours. Detailed live data were received for each patient for 69.5 (53.3-82.8) hours, providing more than 217,000 paired data points on SpO_2_ and oxygen flow per patient during the automated oxygen period, Table 2.

**Table 2.**
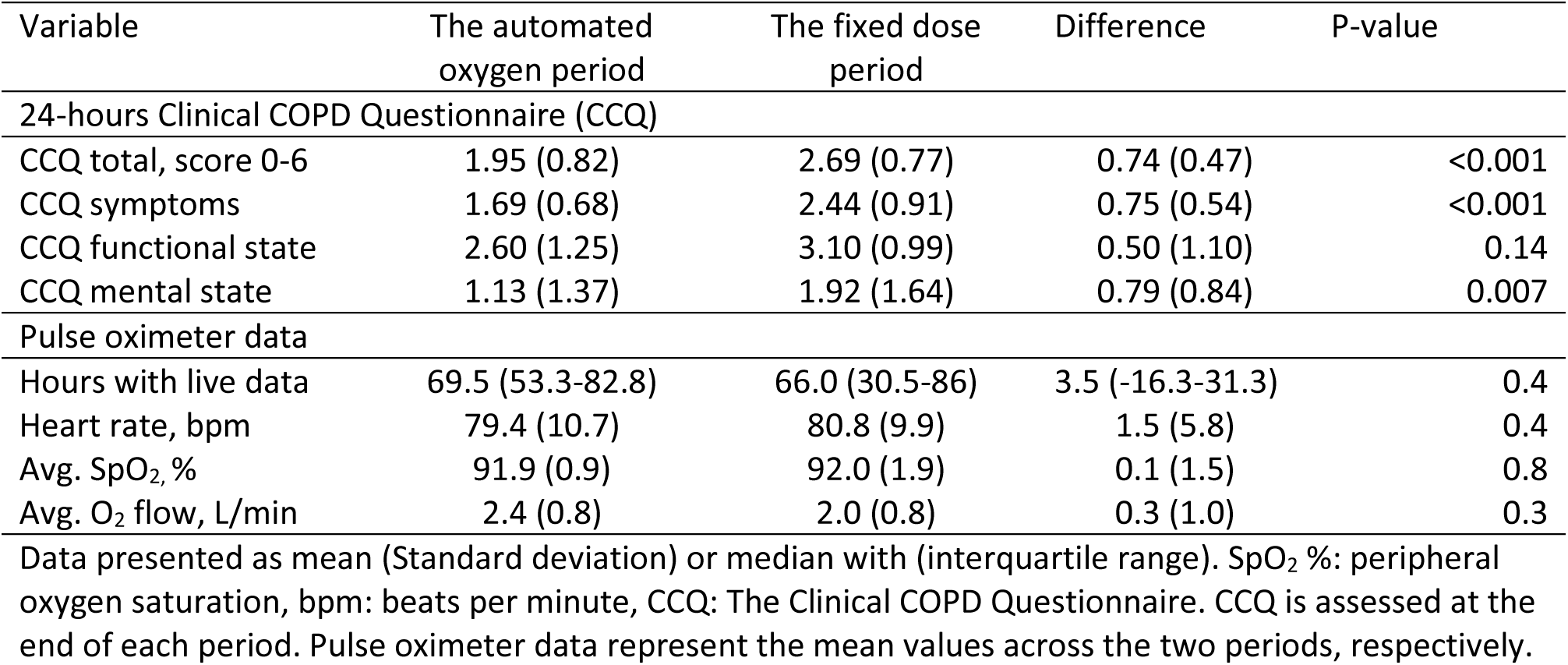
Patient reported outcome and physiological parameters, n=12.

Criteria 1, *Titration time*: Data were received on the online platform for 77 (68.0–84.3) hours, meaning that patients’ oxygen flow was automatically titrated for 83% (74-88) of the possible time.

Criteria 2, *Patient willingness*: Live data were received for 35.5 (26.3-42.5) hours for each patient, constituting 75% (64-88) of the daytime period. Of 12 patients, 11 patients wore the pulse oximeter for at least 50% of the time in the automated oxygen period.

Criteria 3, *Clinical relevance*: Time spent with SpO₂ of 90–94% differed significantly between periods: 52% (42–63) with fixed oxygen flow versus 86% (75–90) with automated titration (p=0.002). Correspondingly, significant differences in favor of the automated oxygen intervention were also observed in time spent with moderate hypoxemia (p=0.003), severe hypoxemia (p=0.004), and hyperoxemia (p=0.01), as illustrated in Figure 3.

**Figure 3.**
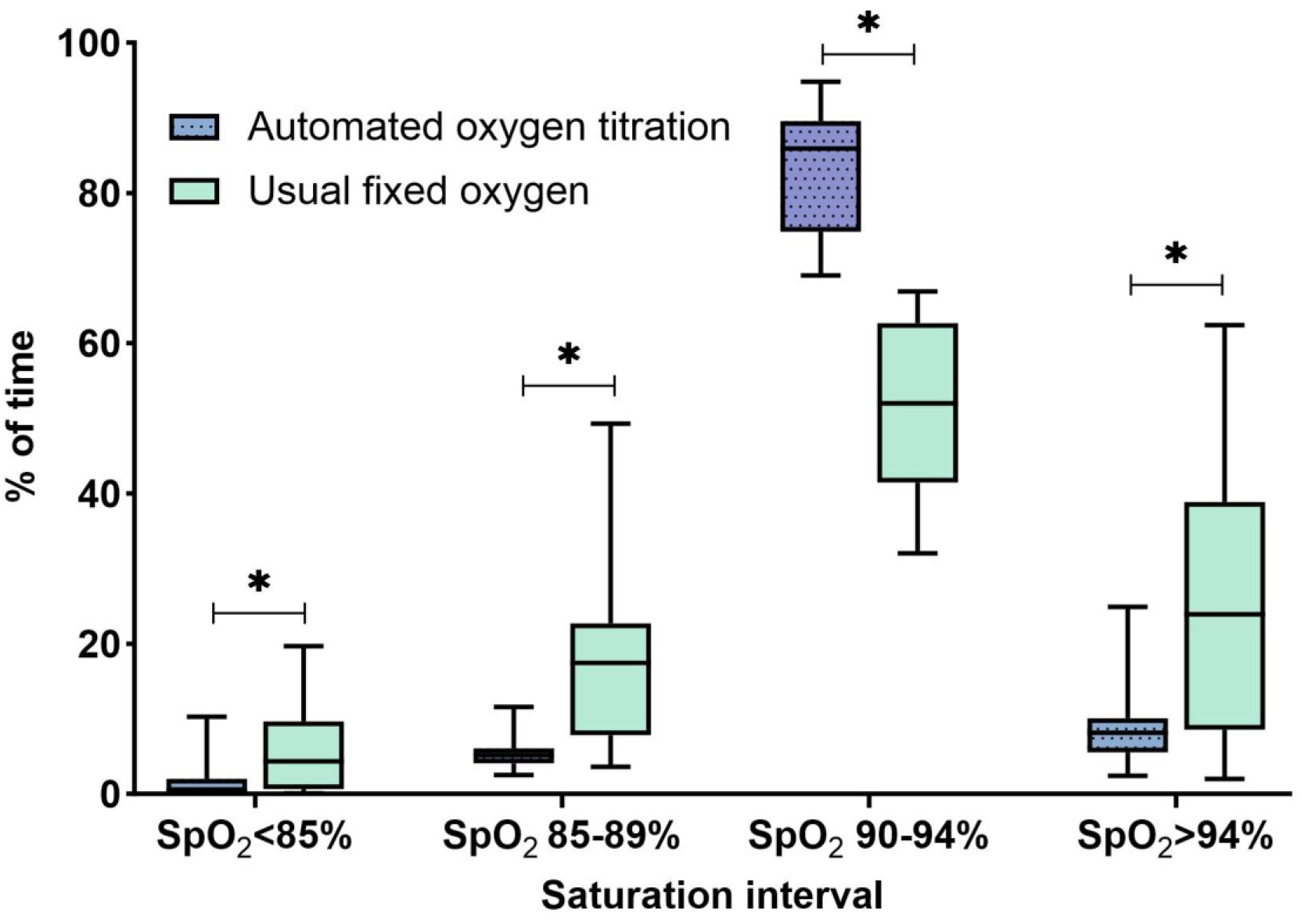
Percentage of time spent within oxygen saturation (SpO_2_) intervals. Boxplot illustrating median, interquartile range, minimum and maximum time spent in the various intervals. X-axis: Four predefined oxygen saturations intervals. Y-axis: Percentage of time over a total period of four full days. *p ≤0.01.

Criteria 4, *Clinical relevance*: The patients took 2297 (1519-3106) steps daily in the fixed dose period and 2366 (1236-3703) steps during the automated oxygen period, with no significant difference between periods (p=0.7).

Criteria 5, *Patient safety:* Unintended events were observed during the intervention; none which required hospitalization or unscheduled healthcare visits. Some events triggered system alarms, primarily due to insufficient oxygen supply, see Table 3. The patients expressed annoyance with the noise from the concentrator, the bulkiness of the pulse oximeter, and the frequent use of batteries.

**Table 3.**
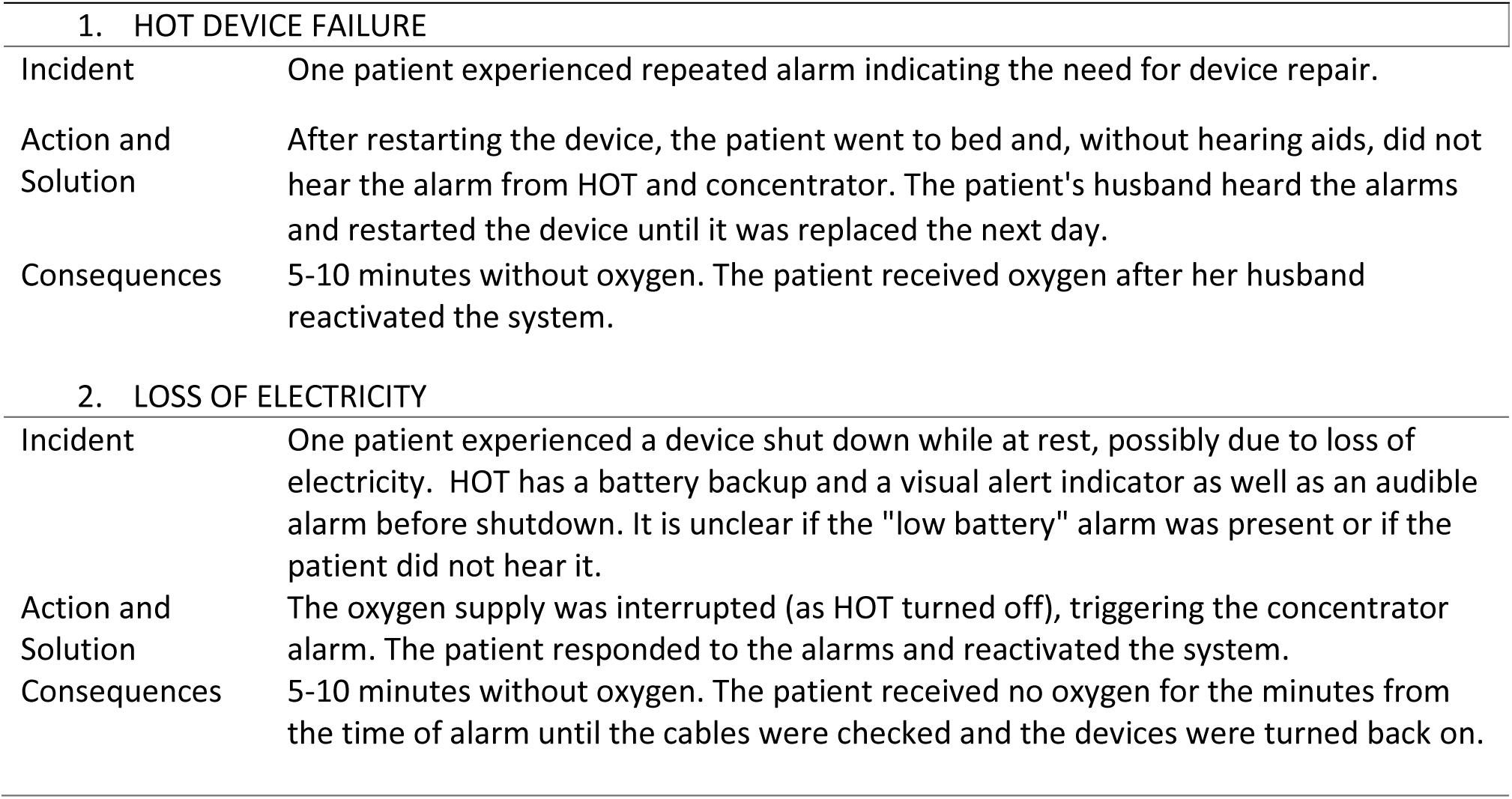
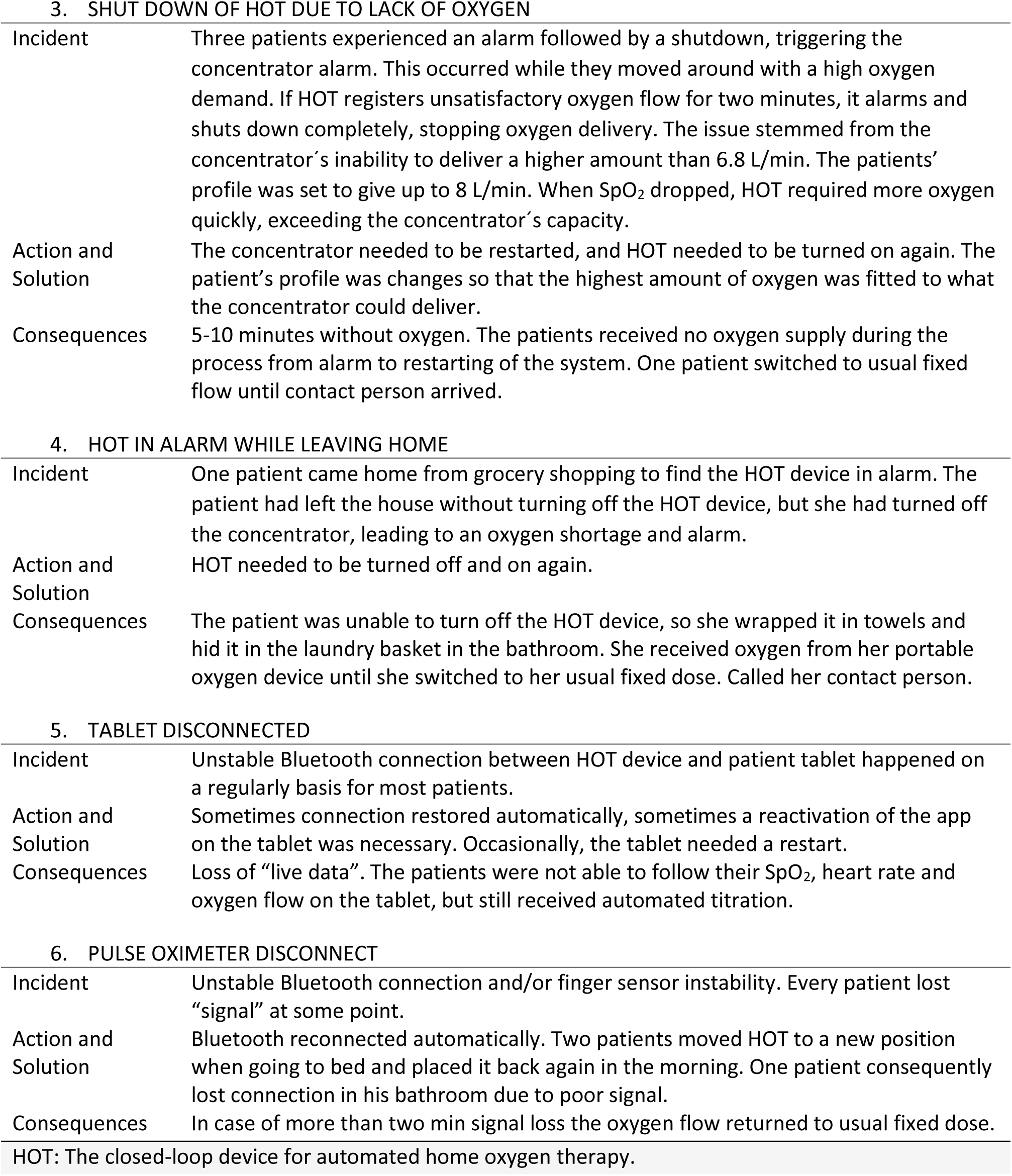
Outlining of incidents during the automated oxygen period.

|  |  |
| --- | --- |
| **1. HOT DEVICE FAILURE** |  |
| Incident | One patient experienced repeated alarm indicating the need for device repair. |
| Action and Solution | After restarting the device, the patient went to bed and, without hearing aids, did not hear the alarm from HOT and concentrator. The patient's husband heard the alarms and restarted the device until it was replaced the next day. |
| Consequences | 5-10 minutes without oxygen. The patient received oxygen after her husband reactivated the system. |
| **2. LOSS OF ELECTRICITY** |  |
| Incident | One patient experienced a device shut down while at rest, possibly due to loss of electricity. HOT has a battery backup and a visual alert indicator as well as an audible alarm before shutdown. It is unclear if the "low battery" alarm was present or if the patient did not hear it. |
| Action and Solution | The oxygen supply was interrupted (as HOT turned off), triggering the concentrator alarm. The patient responded to the alarms and reactivated the system. |
| Consequences | 5-10 minutes without oxygen. The patient received no oxygen for the minutes from the time of alarm until the cables were checked and the devices were turned back on. |
| 3. SHUT DOWN OF HOT DUE TO LACK OF OXYGEN |  |
| Incident | Three patients experienced an alarm followed by a shutdown, triggering the concentrator alarm. This occurred while they moved around with a high oxygen demand. If HOT registers unsatisfactory oxygen flow for two minutes, it alarms and shuts down completely, stopping oxygen delivery. The issue stemmed from the concentrator's inability to deliver a higher amount than 6.8 L/min. The patients' profile was set to give up to 8 L/min. When SpO<sub>2</sub> dropped, HOT required more oxygen quickly, exceeding the concentrator's capacity. |
| Action and Solution | The concentrator needed to be restarted, and HOT needed to be turned on again. The patient's profile was changed so that the highest amount of oxygen was fitted to what the concentrator could deliver. |
| Consequences | 5-10 minutes without oxygen. The patients received no oxygen supply during the process from alarm to restarting of the system. One patient switched to usual fixed flow until contact person arrived. |
| 4. HOT IN ALARM WHILE LEAVING HOME |  |
| Incident | One patient came home from grocery shopping to find the HOT device in alarm. The patient had left the house without turning off the HOT device, but she had turned off the concentrator, leading to an oxygen shortage and alarm. |
| Action and Solution | HOT needed to be turned off and on again. |
| Consequences | The patient was unable to turn off the HOT device, so she wrapped it in towels and hid it in the laundry basket in the bathroom. She received oxygen from her portable oxygen device until she switched to her usual fixed dose. Called her contact person. |
| 5. TABLET DISCONNECTED |  |
| Incident | Unstable Bluetooth connection between HOT device and patient tablet happened on a regularly basis for most patients. |
| Action and Solution | Sometimes connection restored automatically, sometimes a reactivation of the app on the tablet was necessary. Occasionally, the tablet needed a restart. |
| Consequences | Loss of "live data". The patients were not able to follow their SpO<sub>2</sub>, heart rate and oxygen flow on the tablet, but still received automated titration. |
| 6. PULSE OXIMETER DISCONNECT |  |
| Incident | Unstable Bluetooth connection and/or finger sensor instability. Every patient lost "signal" at some point. |
| Action and Solution | Bluetooth reconnected automatically. Two patients moved HOT to a new position when going to bed and placed it back again in the morning. One patient consequently lost connection in his bathroom due to poor signal. |
| Consequences | In case of more than two min signal loss the oxygen flow returned to usual fixed dose. |

HOT: The closed-loop device for automated home oxygen therapy.

### Secondary outcome

The total score in the CCQ improved significantly by a mean of 0.74 (0.47) points favoring the automated oxygen intervention (p<0.001), Table 2.

No difference in mean oxygen flow between periods was observed (p>0.3), Table 2. However, all 12 patients used the full range of possible oxygen flow during the automated oxygen period. During 34% (18) of the measured time, the patients’ oxygen flow was titrated up compared to their usual fixed oxygen dose, and in 38% (27) of the time it was decreased, Figure 4.

**Figure 4.**
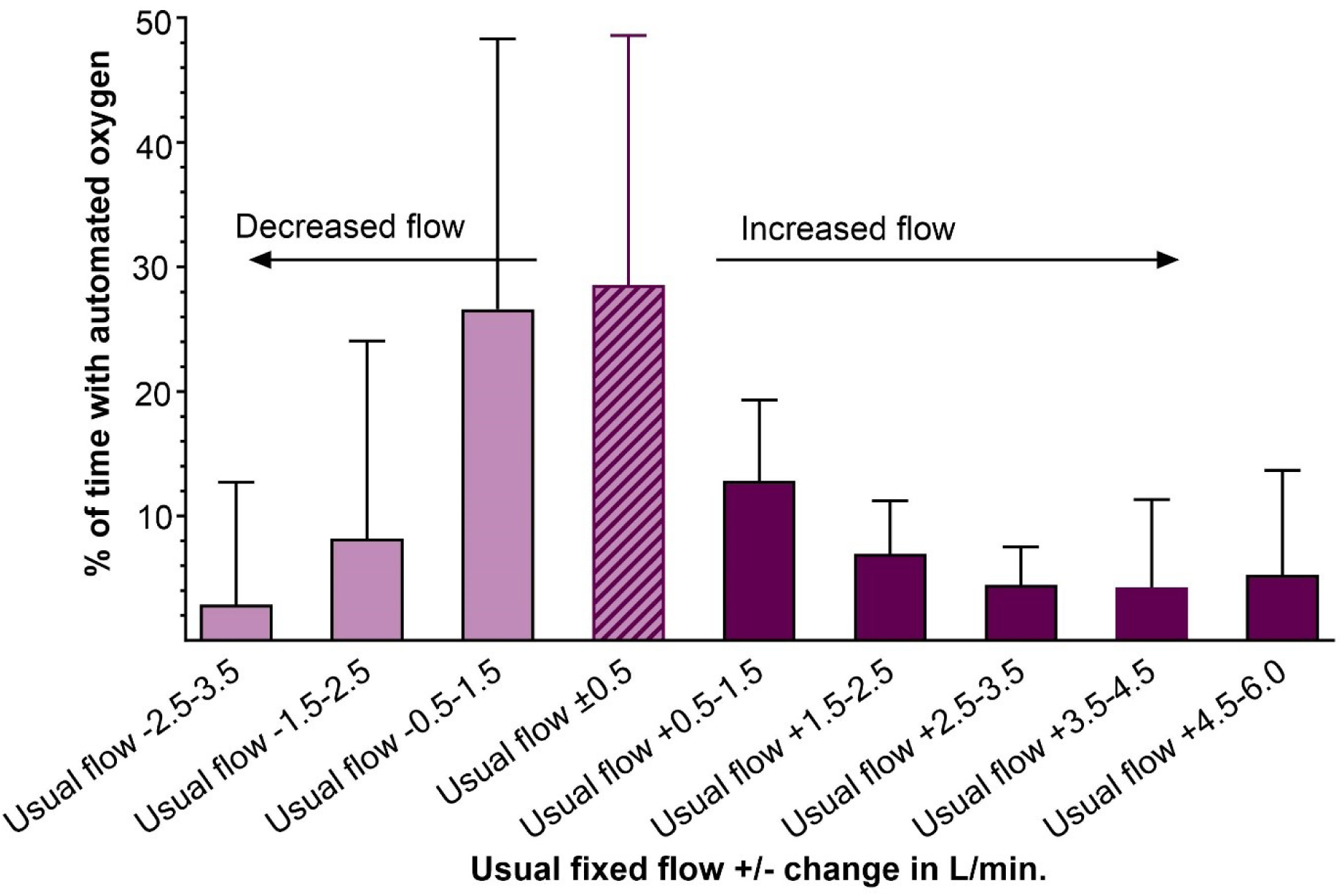
Various oxygen flow used during the automated oxygen period. The percentage of the time the patients spent using various oxygen flows during automated titration, expressed as changes in 1 L/min intervals from their usual fixed flow ±0.5 L/min. The striped bar represents the 29% (20) of the time, that the patients received their usual oxygen flow. The light purple bars indicate the percentage of time patients experienced decreased oxygen flow compared to their usual flow, while the dark purple bars represent the time in which the oxygen flow was increased.

The test for carryover-effect showed neither significant difference in CCQ total score between the two periods nor significance of an interaction term between treatment and the period.

## Discussion

We found that continuous automated oxygen titration in the homes of patients with COPD on LTOT was both feasible and well-tolerated, with high patient compliance. For more than 80% of the day, the patients’ oxygen flow was titrated based on their saturation, resulting in a significant improvement in oxygenation at home compared to when using their fixed dose. Furthermore, the patients reported a lower symptom burden, as measured by the CCQ, indicating a more positive perception of health status when oxygen saturation was improved.

### Target saturation and oxygen flow

Interestingly, the mean oxygen flow and mean saturation levels were nearly identical across both study periods, which could suggest that automated oxygen titration offers limited added value. However, in 71% of the time that patients used automated oxygen delivery, their oxygen flow was automatically adjusted to a different rate than their usual fixed dose. Not only did the oxygen flow increase with up to additional 5 L/min, but in more than 33% of the time, the patientś oxygen flow was decreased compared to their usual fixed dose. This highlights the fluctuating oxygen needs at an individual level for each patient. This variability in oxygen flow corresponded to the significant improvements in the time spent within the target saturation. In line with these findings, fluctuating oxygen needs have been observed during walking [17–19], as well as in more home-related settings, such as during an ADL test. In this context, the mean oxygen flow increased from 1.6 to 5.2 L/min, while the time spent with severe hypoxemia (SpO2 <85%) decreased by 30% to 17% [20]. These improvements in oxygen saturation led to enhanced performance and reduced experiences of dyspnea.

We found that the patients spent approximately 22% of the day with SpO₂ below 90% while using their usual fixed oxygen flow. This is slightly different from a study by Sliwinski et al., in which patients spent 30% of the day with hypoxemia [11]. Patients are typically more active outdoors, thus probably also more frequently experiencing hypoxemia, and this could explain the differences in time spent with hypoxemia between the present study and Sliwinski’s, where patients also were monitored while outdoors.

Sliwinski did not report how long patients maintained a saturation above 94%. However, Zhu et al. argued that oxygen flow rates should be reduced during rest, as patients’ oxygen saturation were often too high [15]. In present study, we were surprised to find that patients with usual oxygen dose spent 24% of their time with saturation above the target range while at home. This corresponds to six hours per day with hyperoxemia, during which the patients could use a lower oxygen flow or, depending on the degree of respiratory failure, take a break from the nasal cannula. In line with this, Ekström et al. recently found that patients could pause oxygen therapy for up to nine hours per day without any consequences on mortality or hospitalization [25]. However, they did not monitor for how long time the patients were well treated without hypoxemia [25]. The NOTT and MRC studies from the 1980s established that reducing hypoxemia at rest by increasing PaO_2_ above 8 kPa reduces mortality [26,27]. Oxygen use for 24 hours was found to be more beneficial than 12 hours, and 15 hours was better than no oxygen at all. However, it remains unclear whether the critical factor is the overall duration of oxygen therapy, or the time spent with effective oxygen therapy (PaO₂ >8 kPa). While the effect on mortality is one consideration, it is well established that improved oxygenation supports patients’ physical functioning and helps reduce breathlessness in controlled test settings. Whether improved oxygenation could lead to similar benefits on mortality in the home environment remains to be investigated, but our results suggested an effect on the patientś perceived symptoms which could be important to their daily life.

### Health status and daily step counts

Health status, evaluated using the CCQ, could potentially detect improvements in patients’ ability to move with less dyspnea and participate in more social activities. In our 12-patient feasibility study, the CCQ overall score significantly favored the automated oxygen period, with a striking difference of 0.74 point, which is almost the double of the 0.4 MID established for the CCQ in this population. Both the symptoms and mental state domains of the CCQ showed significant improvement, supporting the hypothesis that optimized oxygenation can alleviate dyspnea, as also observed during walking and in ADL. Limited evidence exists on home-based interventions for patients with COPD that effectively improve health-related quality of life, particularly for those with severe disease, who often experience profoundly diminished quality of life [28,29]. Consequently, any home-based oxygen intervention that meaningfully reduces patients’ symptom burden is highly relevant.

In general, the patients in the present study were not very active with approximately 2,300 steps per day, and with no significant difference between the two groups. Still, this aligns with previously reported for this severely ill group, typically ranging between 2,400 and 3,800 steps per day [30]. Given the short intervention of four days and that the intervention was only indoor in the present study, our criteria for feasibility related to physical activity (no difference between groups) was met. The study was not powered to find any difference in step counts and considering how difficult it is to change habits we did not expect to find a difference between groups.

An essential factor in evaluating feasibility before considering a larger trial or implementation is the patients’ attitude toward the intervention. This includes the perceived burden of the technology and how well it aligns with what patients find personally meaningful and effective. Patients’ experiences are reported in a separate paper, gathered through qualitative interviews with the involved patients in the present study [31].

### Equipment challenges

The proper solution for maintaining a target saturation only emerged with the introduction of automated closed-loop devices for home use. Effective automated oxygen administration, aligned with our study goal of continuous saturation monitoring during daytime followed by automatic oxygen adjustments, requires the optimization of all system components. This includes the closed-loop device, the pulse oximeter, and the oxygen concentrator. Additionally, a patient tablet, strong Wi-Fi and coordination between all mentioned system parts are essential. A well-designed nasal cannula capable of delivering flows above 5 L/min without causing discomfort to the patient is also preferable. Early in the study, it became apparent that the concentrators were unable to sustain an 8 L/min flow, leading us to lower the maximum flow for consistent performance. Furthermore, the standard concentrator, typically used for flow rates higher than 5 L/min, was notable noisy and the reason for the patientś primary complains. The pulse oximeter was by many found to be uncomfortable for constant wear and too demanding in battery use.

Before starting the study, we decided that data loss was important to look at (criteria 1). However, it turned out to be impossible to distinguish between times when the oximeter was deliberately removed (e.g., for a shower or when leaving the home), and time when the Bluetooth connection was unintentionally lost or when equipment malfunctioned. Therefore, we chose to disregard the 10% threshold, as it proved to be an unsuitable target.

Adding an electronic device to the patient’s oxygen therapy system increases the risk of unintended events and system alarms. Some of these alarms are appropriate as they alert to an undesirable situation, such as low oxygen flow. Acting on alarms under current conditions requires that the patients have both auditory and visual awareness, along with sufficient cognitive capacity to respond adequately to alarms or, at a minimum, switch to their standard treatment if needed.

### Strength and limitation

A key strength of the present study is the home setting, where patients behaved as they typically would, without the need for round-the-clock support. They engaged in their usual daily activities and moved around with their normal level of physical activity. This provided valuable insight into how automated oxygen titration might function in real-world conditions and its potential impact on patients’ daily lives. We believe, the data we received were reflective of real-life scenarios.

Another strength is the extensive amount of data collected for each patient. We collected data over two four-day periods, providing a detailed overview of time spent in different SpO₂ intervals with and without the automated oxygen delivery. The same wrist pulse oximeter ensured consistent saturation measurements during both periods.

A limitation of our study was the non-blinded design, which might lead patients to overestimate the positive impact of more adequate oxygenation. Additionally, the possibility for patients to contact the principal investigator in case of problems might have provided a comforting effect, making patients feel safer and more relaxed, despite this option being available in both study arms. Furthermore, the sample size is small in this feasibility study design, which limits our ability to draw definitive conclusions about the potential effects of the automated oxygen intervention.

### Conclusion

Conclusively, automated oxygen therapy in the patientś home was feasible and it significantly increased the time patients spend with normoxia. This improvement seemed to potentially lower symptom burden such as dyspnea in daily life. To ensure safety and for patients to respond adequately to alarms, the ideal patient must have cognitive awareness as well as adequate hearing and vision.

## Data Availability

Public deposition of raw data points is not possible due to Denmarks national legislation (Data Protection Act section 10 and Data Disclosure Proclamation Act) which outline that we can only transfer pseudonymized data to the Journal after the Data Protection Authorities approval (Data Protection Act section 10, part 3, nr. 3.).
Reviewers and others may obtain access to the data by request, and after the Danish Data Protection Agency has approved of the data transfer from the Capital Region. If others are to gain access to the pseudonymized data, they shall ensure that is an adequate legal basis to share the Capital Regions data and ensure that the data is only being processed for scientific research purposes.

## Acknowledgement

A special thank you to Ida Heefelt for her support and assistance with blood gases. Thanks to O2matic Ltd. for adjusting the equipment to align with our study requirements and for consistently incorporating feedback from both our study team and the patients.

## Declaration of generative AI and AI-assisted technologies in the writing process

During the preparation of this work the first author, used ChatGPT in the writing process in order to improve the readability of the manuscript. After using this tool/service, the authors reviewed and edited the content as needed and take full responsibility for the content of the published article.

